# Multiomic analyses uncover immunological signatures in kidney transplantation

**DOI:** 10.1101/2024.07.15.24309961

**Authors:** Claire Tinel, Alexis Varin, Dany Anglicheau, Jasper Callemeyn, Jetty De Loor, Wilfried Gwinner, Pierre Marquet, Marion Rabant, Virginia Sauvaget, Elisabet Van Loon, Baptiste Lamarthée, Maarten Naesens

**Author notes:** These authors contributed equally (Baptiste Lamarthée, Maarten Naesens). Corresponding author: Baptiste Lamarthée, PhD Rue du Docteur Girod, 25000 Besançon, France.

## Abstract

Identifying biomarkers in kidney transplant patients is essential for early detection of rejection, personalized treatment and improved overall outcomes. It improves our ability to monitor the health of the transplanted organ and tailor interventions to the specific needs of each patient. Here we compiled a multicenter, multiomic dataset of the kidney transplant landscape. Using multi-omics factor analysis (MOFA), we sought to uncover sources of biological variability in patients’ blood, urine and allograft at the epigenetic and transcriptomic levels. MOFA reveals multicellular immune signatures characterized by distinct monocyte, natural killer and T cell substates explaining a large proportion of inter-patient variance. We also identified specific factors that reflect allograft rejection, complement activation or induction treatment. Factor 1 mainly explained the molecular variations in patients’ circulation and discriminated antibody-mediated rejection from T-cell mediated rejection. Factor 2 captured some of the molecular variation occurring within the allograft and associated with complement/monocytes crosstalk. Factor 4 captured the impact of ATG induction. These data provide proof-of-concept of MOFA’s ability to reveal multicellular immune profiles in kidney transplantation, opening up new directions for mechanistic, biomarker and therapeutic studies.

## Introduction

Kidney transplantation is the treatment of choice for patients with end-stage renal disease, offering a significant improvement in quality of life and a reduction in morbidity and mortality. However, the long-term success of renal transplantation depends on many factors, such as immunological compatibility between donor and recipient, optimal management of immunosuppressive therapies and early detection of any signs of rejection or post-transplant complications. In kidney transplantation, both immune and non-immune mechanisms contribute to the progression of histological lesions and scarring of the kidney graft. These lesions, resulting from complex interactions between immune cells, soluble molecules and graft architecture, compromise graft function and long-term survival.

The objectives of the BIOMARGIN study were to discover biomarkers of renal allograft lesions derived from biopsies, blood and/or urine. To achieve these objectives, the BIOMARGIN consortium employed an ambitious strategy involving a succession of clinical studies in 4 hospitals in three European countries for the discovery, confirmation and validation of biomarkers in which transplant patients provided blood, urine and biopsy samples. Large-scale supervised explorations, also known as “omics”, of these biological samples separately using state-of-the-art analytical technologies led to the discovery of several biomarkers or insights in kidney transplantation ^1–5^.

In addition, recent advances in omics technologies have led to unprecedented efforts to characterize the molecular changes underlying the pathophysiology of a wide range of complex human diseases such as coronary syndrome recently^6^. The combination of different omics technologies, called “multi-omics” analyses, has been proposed to decipher the molecular mechanisms involved in complex diseases. These analyses can be classified into supervised and unsupervised methods. The aim of supervised methods is to predict one or more conditions related to a sample, although over-fitting can be an issue. In contrast, unsupervised methods explore the data by analyzing correlations between samples in order to condense the large volume of data into a reduced number of factors which, in turn, could be associated with clinical features. MOFA2 (Multiple Omics Factor Analysis version 2) is an unsupervised statistical approach developed to explore and integrate multiple omics data sources such as genomics, transcriptomics, proteomics and metabolomics^7^. In the present report, we used MOFA2 to uncover specific biological signatures associated with kidney transplant phenotypes and outcomes by identifying complex patterns and relationships between different molecular variables such as mRNA and miRNA from biopsies, blood and urine.

## Results

### MOFA application on BIOMARGIN datasets

We collected data from blood, biopsy and urine samples of 131 kidney transplant recipients comprising six data types (also called views): 1 blood-derived epigenome (miRNA expression), 2 blood-derived transcriptomes (mRNA quantified by MicroArray, mRNA quantified by RNA sequencing), 1 biopsy-derived epigenome (miRNA expression), 1 biopsy-derived transcriptome (mRNA quantified by MicroArray) and 1 urine-derived selected gene set (mRNA quantified by RT-qPCR). In blood, a total of 58828 genes were measured by RNAseq, 54675 genes were detected by MicroArray and 758 miRNAs were measured by RT-qPCR. In biopsy, 54613 genes were assessed by MicroArray and 758 miRNAs were measured by RT-qPCR. In urine, 34 genes were measured by RT-qPCR (**Figure 1a**). Given these 6 data matrices with measurements of multiple omics data types across sample sets or partially overlapping sample sets, MOFA infers an interpretable low-dimensional data representation of factors. These learned factors capture the main sources of variation between views, facilitating the unsupervised identification of continuous molecular gradients or discrete sample subgroups (**Figure 1a**). In order to integrate the various omics data with MOFA, we constructed 131 multiomics profiles by matching samples in the 6 views. It should be noted that only 26.7% (N=35) of the 131 samples were profiled with all types of omics data mainly due to the limited number of samples with mRNA data for urine (**Figure 1b**), but such a scenario of missing values is not uncommon in multidimensional cohort studies and MOFA is designed to cope with it^7^.

**Figure 1:**
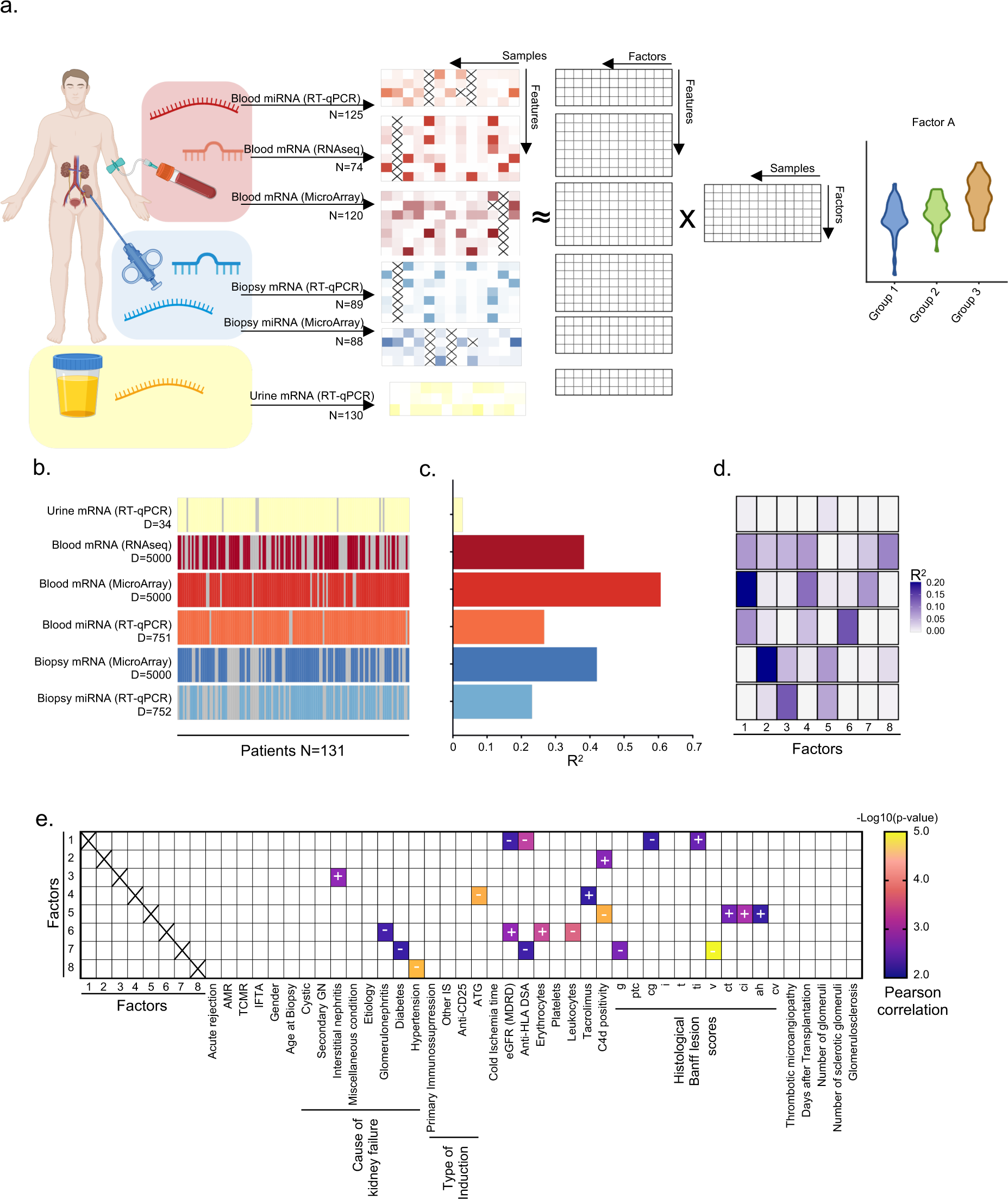
Multi-Omics Factor Analysis application on BIOMARGIN datasets identifies important clinical markers in kidney transplantation. (a) Graphical representation of MOFA’s matrix decomposition of each view’s data into a product composed of a view-specific factor loadings matrix and a shared latent factor matrix. The number of samples (’N’) used for multi-omics factor analysis per data set is indicated. The loading of a given factor can be compared a posteriori between patient groups. (b) Number of features (”D”) in each view. A grey bar indicates that the sample is missing in the given omic view (c) Total percentage of variance explained (R2) per omic view (d) Percentage of variance explained by each latent factor in the different omic views for the selected MOFA model. (e) Pearson correlation matrix analysis of the 8 latent factors and the various clinical parameters indicated. Positive (+) or negative (-) correlations with the loading of each latent factor are indicated, as well as the log10 p-value resulting from the Pearson correlation tests. Panel a was created using Biorender.com.

As it is recommended to perform a stringent selection of features before creating the MOFA object, we performed an initial selection of features from the Blood RNA-seq, Blood miRNAs and Biopsy miRNAs, keeping only the top 25% of genes with the greatest variance (First features selection). Integrating these features with MOFA resulted in an over-representation of transcriptome views at the expense of epigenome views (**Figure S1a**). Secondly, to ensure that epigenome views were not under-represented when fitting MOFA, we removed weakly expressed features from transcriptomic views and filtered out the least expressed genes (Second features selection). This selection resulted in a more balanced representation of blood-derived miRNAs, but did not improve the representation of biopsy-derived miRNAs (% explained variance < 5%, **Figure S1a**). Thirdly, we followed a strategy recommended by the MOFA authors to adjust the number of transcriptome features, selecting the 5000 genes with the greatest variability measured by standard deviation but also taking into account all epigenomic features. We observed that estimating MOFA with a greater number of epigenomic features led to a significant increase in the percentage of variance explained in the epigenome and to a more balanced final features selection. (**Figure S1a and Figure 1c**).

To obtain a MOFA model with this final features selection, we trained MOFA 100 times. We chose the MOFA model with the lowest absolute Evidence Lower Bound (ELBO) value to strike a balance between model complexity and explanatory power (**Figure S1b**). MOFA identified 8 factors that were largely orthogonal, capturing independent sources of variation. Cumulatively, the 8 factors explained 38% of the variation in blood-RNAseq data, 61% in blood-MicroArray data, 27% in blood-epigenome data, 42% in biopsy-transcriptome data, 23% in biopsy-epigenome data and 2.9% in urine data (**Figure 1c**). Of these, Factor 2 and Factor 3 were active in most assays (**Figure 1d**), indicating extensive roles in multiple molecular layers in both the transplanted kidney and the circulation. In addition, other factors such as Factor 1 and Factor 4 were specific to blood-related views. The fact that most of the factors explain the variance between several views indicates that the data are very highly correlated between views, and that it is possible to identify common patterns between different omics views. In contrast, Factor 6, Factor 7 and Factor 8 were mainly active in only one data modality which diminishes their interest in this multiomics approach (**Figure 1d**). We next estimated the correlation between factors and observed correlations R^2^ below 0.4 indicating that each factor captures independent and unique sources of variations (**Figure S1c**).

### MOFA identifies important clinical markers in kidney transplantation

To understand how factors relate to kidney transplant phenotypes, we assessed the relationship between factor loadings and different kidney transplant outcome parameters (**Figure 1e)**. Patient characteristics are shown in **Supplemental Table 2**. Intriguingly, some factors correlated exclusively with Banff histological lesions or complement activation: Factor 2 was positively correlated with C4d deposition in peritubular capillaries (“C4d positivity”; P value<0.001) while Factor 5 was associated with C4d positivity (R^2^<0, P value<0.0001), tubular atrophy (“ct”; R^2^>0, P value<0.01), interstitial fibrosis (“ci”; R^2^>0, P value<0.001) and arteriolar hyalinosis (“ah”; R^2^>0, P value<0.01). In contrast, Factor 3 and Factor 8 correlated uniquely with certain causes of renal failure (interstitial nephritis, R^2^>0, P value<0.001 and hypertension, R^2^<0, P value <0.0001 respectively). Factor 4 was strongly associated with immunosuppression: it was negatively correlated with ATG induction (P value <0.0001) and positively correlated with tacrolimus immunosuppression (P value <0.01). In addition, certain factors were associated with multiple parameters of varying natures. Factor 7 was negatively associated with diabetes as a cause of renal failure (P value<0.01), creatinine (P value<0.0001), anti-HLA DSA (P value<0.01), g lesions (P value<0.01) and v lesions (P value<0.00001). Factor 1 correlated with eGFR (R^2^<0, Pearson correlation P value<0.01), anti-HLA DSA (R^2^<0, P value<0.001), transplant glomerulopathy (“cg”; R^2^<0, P value<0.01) and total inflammation (“ti”; R^2^>0, P value<0.01). Lastly, Factor 6 was associated with glomerulonephritis as a cause of kidney failure (R^2^>0, Pearson correlation P value<0.01), eGFR (R^2^>0, P value<0.01), erythrocyte titer (R^2^>0, P value<0.001), leukocyte titer (R^2^<0, P value<0.001) and ti (R^2^>0, P value<0.01).

### Factor 1 discriminates antibody-mediated from T-cell mediated rejection

Factor 1 mainly explained the molecular variations in blood of kidney transplant recipients (**Figure 1d**) and was negatively correlated with anti-HLA DSA status. Intriguingly, this blood-related factor was also associated with cg and ti lesions in the graft (**Figure 1e**). To confirm these correlations, we assessed the global distribution of Factor 1 loading (**Figure 2a**), and then explored the distribution of Factor 1 loading according to cg and ti lesion scores. We observed a significant decrease in Factor 1 loading in cases with a cg>0 score (**Figure 2b**), while Factor 1 loading increased significantly in cases with a ti>1 score (**Figure 2c**). We also confirmed the significant decrease in Factor 1 loading in the presence of anti-HLA DSA (**Figure 2d, Supplemental Table 3**). We also performed univariate logistic regressions and reported the performance of Factor 1 to discriminate HLA-DSA status and the presence of ti lesions and cg lesions by (**Figure S2a**). Given the increase in Factor 1 loading with ti lesions and the decreases in the presence of HLA-DSA and cg lesions, we tested whether Factor 1 loading differed between cases with antibody-mediated rejection (AMR) and T-cell mediated rejection (TCMR). To this end, we stratified patients according to different phenotypes (AMR, TCMR, Normal and IFTA) and we observed a significant difference between the TCMR and AMR group suggesting that Factor 1 may discriminate TCMR from AMR (**Figure 2e**).

**Figure 2:**
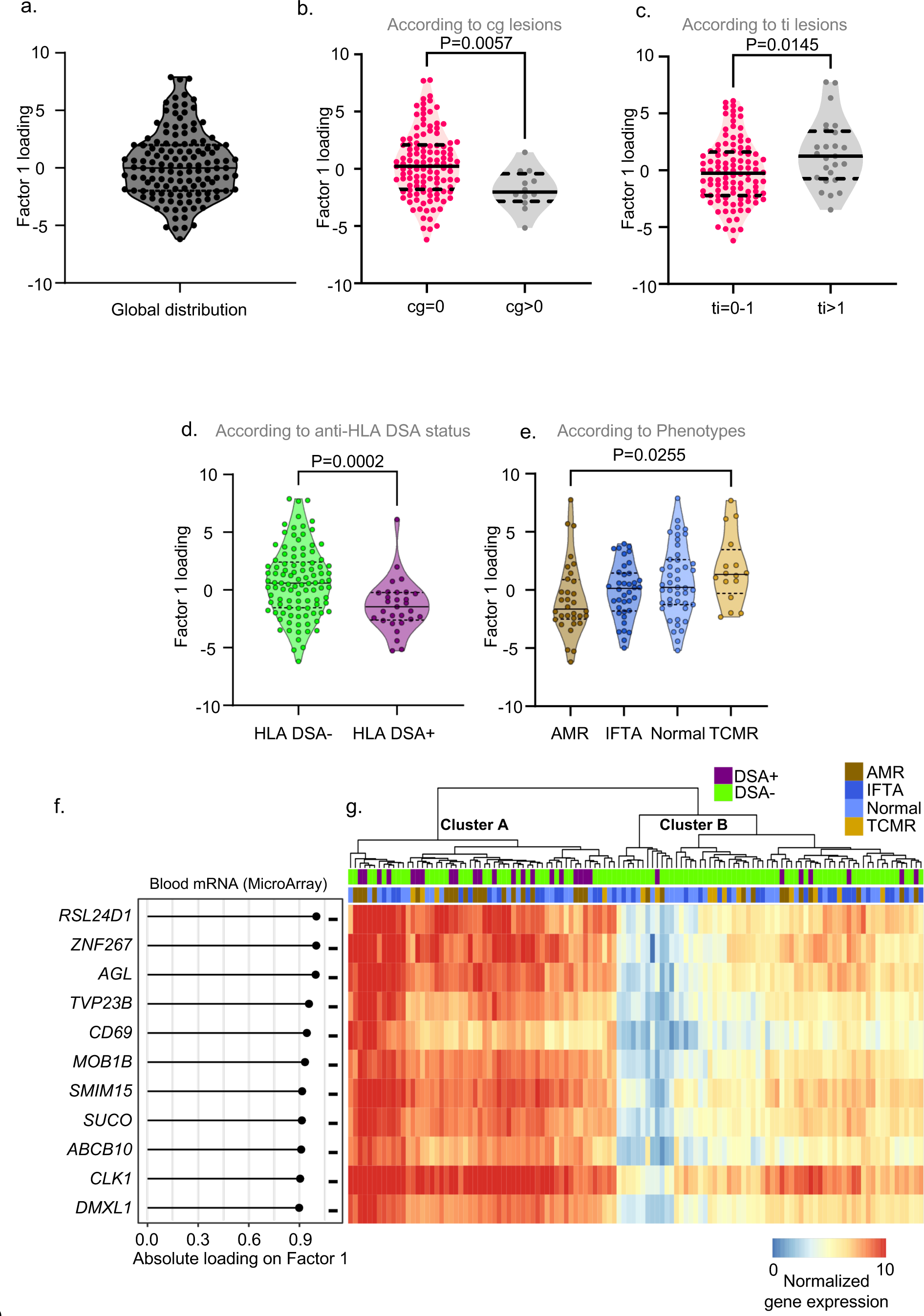
Factor 1 discriminates antibody-mediated from T-cell mediated rejection. (a-e) Violin plots representing the distribution of latent Factor 1 (a) Global distribution (b) According to Banff cg histological lesions. The difference between groups was assessed by a two-tailed Mann-Whitney test (c) According to Banff ti histological lesions. The difference between groups was assessed by a two-tailed Mann-Whitney test (d) According to HLA status. The difference between groups was assessed by a two-tailed Mann-Whitney test (e) According to phenotypes. The difference between groups was assessed by an ordinary one-way ANOVA test and multiple comparisons using the Tukey’s test. (f) Lollipop graph shows the top-weighted genes derived from the MicroArray blood dataset in latent Factor 1. (g) Heatmap showing the distribution of the top-weighted genes in the samples. A and B clusters A were determined here in an unsupervised manner.

Features contributing more than 0.9 of the absolute Factor 1 loading mainly derived from blood MicroArray data (D= 11, **Figure 2f**). The other molecules contributing to each view have also been represented in **Figure S2b**. The significant contribution of the blood MicroArray data view to the Factor 1 is reflected in the heatmap, where a clear separation was observed between blood samples (**Figure 2g**). In fact, unsupervised clustering of blood samples using these 11 top features resulted in two main clusters: samples with HLA-DSA were preferentially grouped in Cluster A in which the 11 features were overexpressed (two-tailed Fisher’s exact test, P-value = 0.001). For the grouping of cases according to AMR or TCMR phenotypes, the Fisher exact test P-values were 0.0562 and 0.1676 respectively. We then used a publicly available blood RNASeq dataset GSE120649^8^ (**Figure S2c**) to validate whether these 11 features in blood MicroArray and the 3 features contributing more than 0.9 of the absolute Factor 1 loading in blood RNA-Seq would be differentially expressed in AMR and TCMR. Interestingly, 12/14 features were found increased in AMR in this dataset (**Figure S2d**), suggesting that Factor 1 could reflect the different immune responses of the two types of rejection in circulating cells. In order to map at the single-cell level the different renal and/or immune cells that could express the top features of Factor 1, we reintegrated 46 publicly available scRNA-Seq datasets from kidney transplant patients, including transcriptomes from both circulating blood cells (PBMC) and cells derived from kidney biopsies (**Figure S3a**). We were thus able to obtain 150,876 transcriptomes which passed quality check (**Figure S3b**) and, using canonical markers (**Figure S3c**), to identify 23 clusters corresponding to all renal and circulating cell populations (**Figure S4a**). For further granularity, a subset corresponding only to circulating cells was selected and 29 clusters were identified and automatically annotated in an unsupervised manner (**Figure S4b**). We then formed a signature corresponding to the top features explaining Factor 1 in the blood and we observed that the top features of Factor 1 were not centralized in a single immune cell population, but instead scattered across myeloid cells, T, B and NK lymphocytes, and even granulocytes (**Figure S4c**). More specifically, *ZNF267* is preferentially expressed by neutrophils, while *MOB1B* and *TVP23B* are derived from basophils and progenitors. *CD69*, *SUCO, SMIM15* and *AGL* are expressed by lymphocytes, *ABCB10* by monocytes and *DMXL1*, *CLK1* and *RSL24D1* by B lymphocytes and plasmablasts. Factor 1 thus corresponds to a multicellular immune profile that differs between TCMR and AMR in patients’ blood.

### Factor 2 is associated with complement/monocytes crosstalk

In contrast to Factor 1, Factor 2 captured some of the molecular variation occurring inside the allograft. Among all the outcomes tested, it was only positively correlated with C4d (Pearson correlation, P-value<0.001) (**Figure 1e**). Of note, C4d deposition in peritubular capillaries is associated with immune reactions directed against the allograft and are the result of activation of the complement system. To confirm the correlation between C4d positivity and Factor 2, we assessed the global distribution of Factor 2 loading (**Figure 3a**), and then explored the distribution of Factor 2 loading according to C4d positivity. We observed a significant increase in Factor 2 loading in cases with C4d positivity (**Figure 3b**). By stratifying patients according to Factor 2 median, a trend towards an increase in the number of C4d-positive cases was also found in patients above the median (**Supplemental Table 3**). The top features contributing to the absolute Factor 2 loading mainly derived from biopsy MicroArray data (D= 15, **Figure 3c**). The top features deriving from biopsy MicroArray data and explaining Factor 2 clearly separated the samples into two groups (**Figure 3d**) in an unsupervised manner. Cases positive for C4d were grouped showed high expression of the top features *LYZ*, *CALHM6*, *EVI2A*, *CD52*, *IGSF6*, *C1QB*, *EVI2B*, *BCL2A1*, *C1QC*, *C16orf54*, *CSTA*, *CXCL9*, *CTSS*, *CXCL10* and *TYROBP*. Interestingly, two of these features *C1QC* and *C1QB* encode the B and C-chains polypeptide of serum complement subcomponent C1q, which associates with C1r and C1s to yield the first component of the serum complement system. With regard to other top features explaining Factor 2 in biopsies, five miRNAs were detected: miR-150, miR-223, miR-1227, miR-624 and miR-155 (**Figure S5a**). Among the upregulated miRNAs, our group has previously reported that miR-155 is preferentially expressed by monocytes^4^, suggesting that Factor 2 may capture the infiltration of blood monocytes/macrophages into the allograft and the overexpression of complement by these cells. In order to validate the association between these Factor2 top features and C4d deposition in peritubular capillaries in an external dataset, MicroArray data from biopsies of kidney transplant patients with and without C4d were analyzed (**Figure 3e)**. Here, the transcriptomes of 23 C4d-negative biopsies were compared with 16 biopsies showing focal, diffuse or minimal C4d ptc staining (**Figure 3f)**. As shown on the volcano plot, biopsies positive for C4d showed overexpression of both mRNA transcripts and miR-150, miR-223 and miR-155 explaining Factor 2. In the scRNA-Seq dataset, we then formed a signature corresponding to the top features explaining Factor 2 in the kidney and observed that this signature was particularly strong in myeloid cells (**Figure S5b**). Strikingly, macrophages were the main cell population expressing the top features (**Figure S5c**) and more specifically *C1QC* and *C1QB*, suggesting that Factor 2 captured the crosstalk between macrophages and complement activation in the allograft after transplantation.

**Figure 3:**
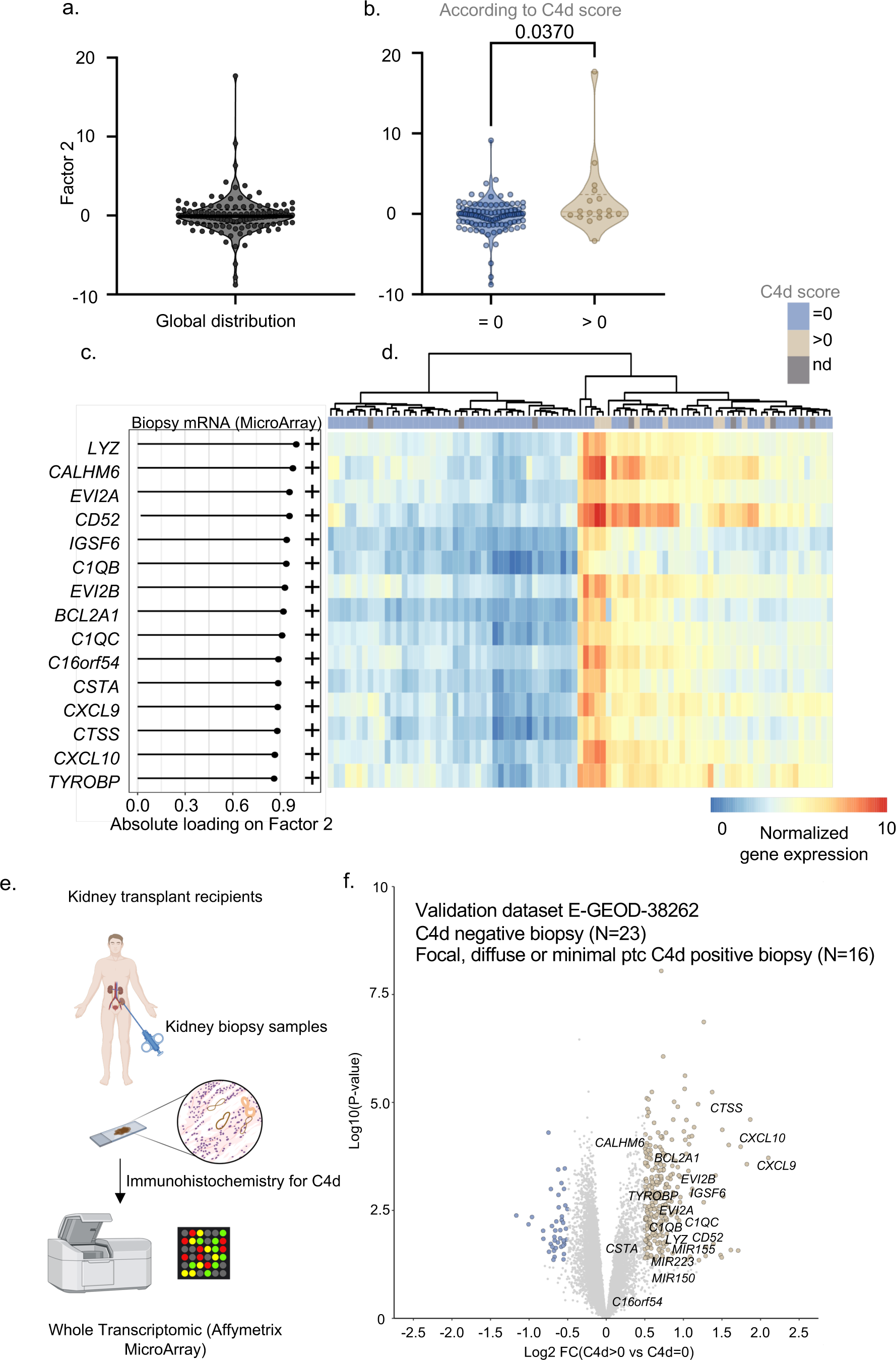
Factor 2 is associated with complement/monocytes crosstalk. (a-b) Violin plots representing the distribution of latent Factor 2 (a) Global distribution (b) According to C4d positivity. The difference between groups was assessed by a two-tailed Mann-Whitney test (c) Lollipop graph shows the top-weighted genes derived from the MicroArray biopsy dataset in latent Factor 2. (d) Heatmap showing the distribution of the top-weighted genes explaining Factor 2 in the samples. (e-f) An external validation dataset E-GEOD-38262 was used to confirm the increase of the top-weighted features explaining Factor 2 in C4d positive biopsies. (e) Experimental scheme (f) Volcano plot showing the differentially expressed genes and miRNAs in C4d positive biopsies (N=16) compared to C4d negative biopsies (N=23). The top-weighted features explaining Factor 2 are indicated. Panel e was created using Biorender.com.

### Factor 4 captures the impact of induction prophylaxis

Similar to Factor 1, Factor 4 captured some of the molecular variation in blood samples (**Figure 1d**). Factor 4 was strongly and negatively correlated with ATG induction (Pearson correlation, P-value<0.00001) and positively correlated with tacrolimus immunosuppression protocol (**Figure 1e**). Evaluating the overall distribution of Factor 4 loading (**Figure 4a**), and then exploring the distribution of Factor 4 loading according to induction types, we observed that Factor 4 loading was also reduced in cases that had received ATG as induction compared to no induction (P-value=0.0023) or other induction therapy (**Figure 4b, Supplemental Table 5**). Considering the top 10 features measured by RNAseq in blood in terms of absolute loading, two clear groups of patients were distinguished in an unsupervised manner: one showing low *IL1R2* expression and without patients who had received ATG (N=0/21, 0%), and a second with high *IL1R2* levels including 15/53 patients who had received ATG as induction (28%). Interestingly, among the other top features, expressions of *TRBC2*, *BCL11B*, *CD3G*, *CD247*, *CD3E*, *STMN3*, *GIMAP7*, *FCMR*, *SRNPN* and *CCR7* were down-regulated in the group including ATG-treated patients (**Figure 4c-d**). The advantage of MOFA is to uncover transcriptomic profiles across omics types and tissues. Strikingly, one of the most weighted features explaining Factor 4 in the biopsy is *IL1R2* (**Figure S6a**).

**Figure 4:**
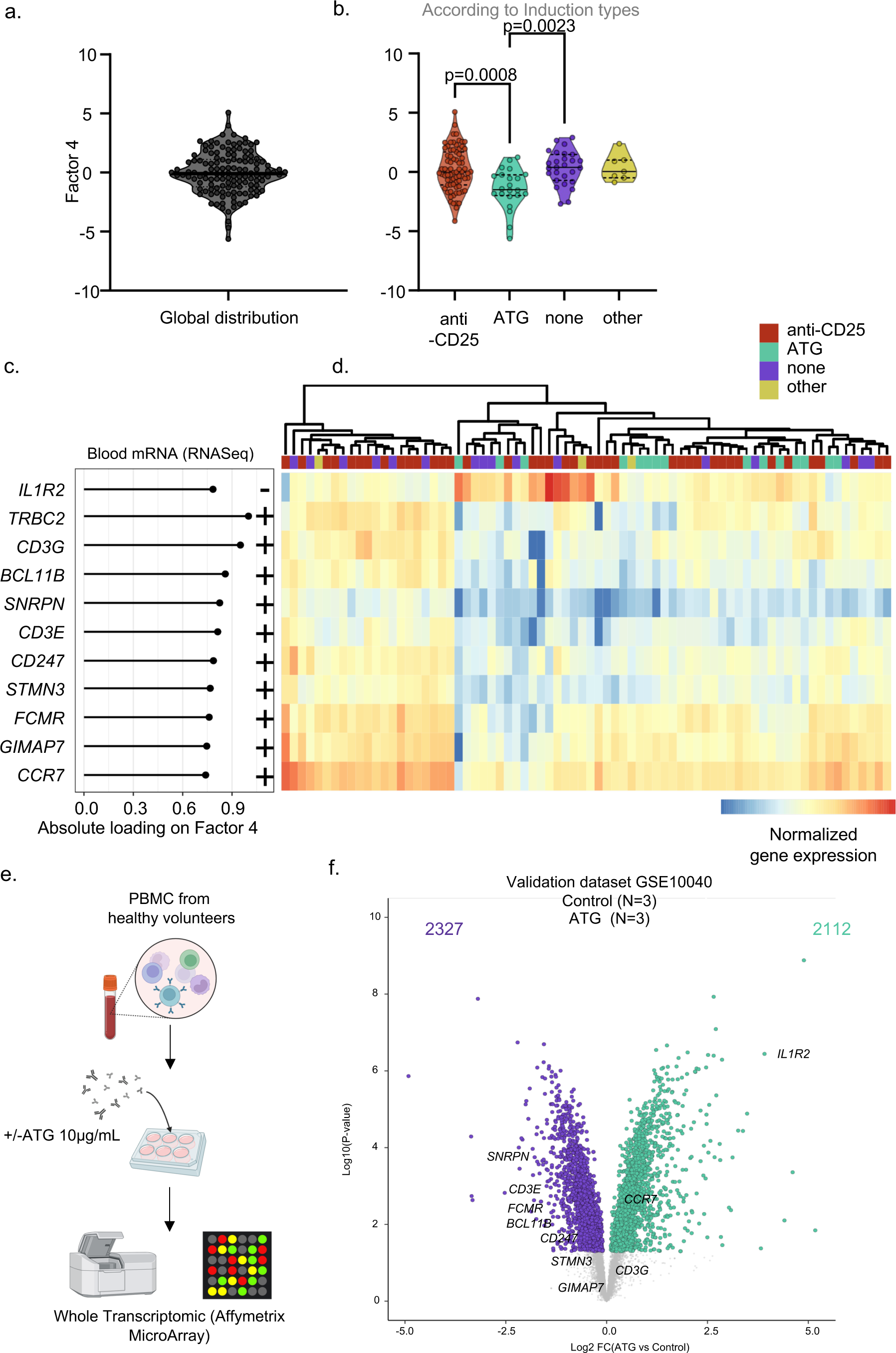
Factor 4 captures the impact of induction prophylaxis. (a-b) Violin plots representing the distribution of latent Factor 4 (a) Global distribution (b) According to induction types. The difference between groups was assessed by a two-tailed Mann-Whitney test (c) Lollipop graph shows the top-weighted genes derived from the RNAseq blood dataset in latent Factor 4. (d) Heatmap showing the distribution of the top-weighted genes of Factor 2 in the samples. (e-f) An external validation dataset GSE10040 was used to confirm the decrease of the top-weighted features explaining Factor 4 in PBMC treated with ATG. e) Experimental scheme f) Volcano plot showing the differentially expressed genes in PBMC treated with ATG (N=3) compared to control (N=3). The top-weighted features explaining Factor 4 are indicated. Panel e was created using Biorender.com. ATG, anti thymo globulin ; PBMC, peripheral blood mononuclear cells

Querying the scRNA-Seq dataset, we observed that the other major features were principally expressed by T cells and NK cells (**Figure S6b**). More specifically, naive T cells subpopulations as well as regulatory T cells and central CD8 T cells highly expressed Factor 4 top features such as *FCMR*, *CCR7*, *SNRPN* and *BCL11B*. *FCMR* expression was mainly restricted to B cells subpopulation. In addition, we observed that *IL1R2* is predominantly expressed by and myeloid dendritic cells and classical monocytes in the blood (**Figure S6c**), suggesting a disturbance in the lymphoid/myeloid cell ratio in both patient groups. To validate these data, we took advantage of a public GSE10040 dataset corresponding to an *in vitro* experiment in which PBMCs from healthy volunteers were treated with ATG at 10μg/mL for 24h^9^. After culture, cells were harvested and mRNA extracted for full transcriptomic analysis (**Figure 4e**). We found that ATG treatment induced the differential expression of 4438 genes (DEGs) comprising 2327 down-regulated genes and 2112 up-regulated genes. By mapping the top features of Factor 4 among these 4438 DEGs, we observed that *IL1R2* is strongly induced by ATG treatment (fold change (FC) >3.9 and Log10(pvalue) >6.4) but so is *CCR7* (FC>0.9, Log10(pvalue) >3.3). In contrast, the other features are mainly down-regulated, such as *SNRPN* (FC<-0.7, Log10(pvalue) >3.5), *CD3E* (FC<-0.3, Log10(pvalue) >1.7), *FCMR* (FC<-0. 9, Log10(pvalue) >2.1), *BCL11B* (FC<-0.5, Log10(pvalue) >1.8), *CD247* (FC<-0.3, Log10(pvalue) >1.3), *STMN3* (FC<-0.1, ns) and *GIMPAP7* (FC<-0.1, ns). Of note, *TRCB2* was not detected and *CD3G* was slightly increased (FC>0.1, ns) (**Figure 4f**). Overall, these results suggest that ATG treatment induces a strong disruption of the immune system that could be detected several months after treatment in blood but also in the allograft, leading to a decrease in the lymphoid compartment in favor of the myeloid compartment.

## Discussion

MOFA2 analysis enables researchers to simultaneously combine and analyze high-throughput data from different biological sources. This method of analysis is proving particularly valuable in the field of medical research, where it enables us to better understand the complexity of the molecular and cellular interactions involved in complex disease processes. By elucidating these mechanisms, clinicians can better target therapies and improve long-term kidney success. To our knowledge, this is the first time this type of MOFA analysis has been conducted in the field of kidney transplantation. Integrating the different omics layers, MOFA2 delimited 8 different factors in an unsupervised manner. As expected, these factors captured independent transcriptomic profiles across blood and renal allografts.

Surprisingly, we observed very few associations between histology and the factors. This indicates that rejection phenotypes (according to the Banff classification), but also rejection severity and phenotype patterns such as rejectionclass are not the main drivers of molecular heterogeneity within allografts and in circulation. In fact, only Factor 1 reflects different profiles between HLA-DSA positive and HLA-DSA negative cases.

Factor 2 was related to the intragraft crosstalk between monocytes/macrophages and complement. It was associated with C4d deposition independent of rejection status, suggesting that this crosstalk is not specific to the AMR process. Indeed, Factor 2 was not significantly correlated with the presence of HLA-DSA, suggesting that this crosstalk between macrophages and complement might be due to non-HLA antibodies which could be detrimental in the context of kidney transplantation^10–12^. In fact, interactions between monocytes/macrophages and complement factors are multiple and complex, contributing significantly to the innate immune responses. Infiltrating monocytes and tissue-resident macrophages can locally synthesize several components of the complement system, including C1, C2, C3, C4, C5, C6, C7, C8, and C9. In particular, we have previously shown that CD163+ macrophages are the main immune cells expressing complement-associated genes such as *C1QA*, *C1QB* and *C1QC* among myeloid cells present in the allograft^13^. In turn, certain complement degradation products, such as the C5a fragment via CD88 or C3a via Toll Like Receptors or C4d via LILRB2 and LILRB3^14^, act as chemokines, attracting monocytes and macrophages and modulating their production of cytokines such as TGF-β1^15^. It is worth noting that urinary *TGFB1* mRNA was the second most important feature (absolute loading>0.95) explaining Factor 2, and one could speculate that this urinary detection of *TGFB1* mRNA could be due to podocyte lesions^16^. At the same time, biopsy-derived miR-155 was also among the key features explaining Factor 2. Intriguingly, this miRNA was closely linked to podocyte apoptosis after exposure to TGF-β1^17^. Altogether our results suggest that Factor 2 reflects the monocytes/macrophages crosstalk with complement within the allograft and that this crosstalk may be associated with podocyte lesions.

Surprisingly, Factor 4 was significantly associated with the type of induction regimen, suggesting that ATG therapy can affect patients’ immune profiles longer term after transplantation. It is clear that ATG makes patients more susceptible to infections, both in the short and long term^18^. This increased sensitivity may persist even after ATG treatment has been discontinued. ATG induction has also been associated with a higher risk of long-term malignancies compared with anti-CD25 induction^19^. The top-weighted genes of Factor 4 suggest an imbalance in the patient’s immune system, with a decrease in the T cell compartment in favor of the myeloid compartment both in the circulation and in the allograft.

This study has several limitations. The datasets used in this study are relatively small, which may limit the ability to capture the full spectrum of variability and complexity present in larger, more diverse populations. Indeed, our study primarily involves Western European centers with a predominantly Caucasian population. This demographic constraint may limit the applicability of our results to other ethnic groups and geographic regions, potentially reducing the overall generalizability of our conclusions. In addition, the analysis in this study is fully dependent on the MOFA2 algorithm. While MOFA2 is a powerful tool for integrative analyses, relying exclusively on this algorithm means that other integrative approaches might yield different factors and insights. This dependence highlights the need for comparative studies using alternative methodologies to validate our findings. Although we conducted systematic validation on external datasets for each factor of insterest, our study lacks in-depth mechanistic studies to confirm the identified factors with greater certainty.

In conclusion, MOFA2 analysis represents a major advance in the integration of omics data to understand kidney transplantation. Our study highlights the significant associations between factors and clinical parameters. This highlights MOFA as an innovative approach to dissect multicellular immune profiles with mechanistic and clinical implications in kidney transplantation. Furthermore, our study, as a globally available resource, provides new targets for large-scale MOFA-based experimental studies and biomarker assays, and helps prioritize new candidate targets for immunomodulatory interventions in kidney transplantation.

## Methods

### Patient population and data collection

The present study is part of the Reclassification using OmiCs integration in KidnEy Transplantation (ROCKET) project, which is based on the BIOMArkers of Renal Graft INjuries (BIOMARGIN) study (ClinicalTrials.gov number NCT02832661). Patients were included prospectively in four European transplant centers between June 2011 and March 2017 (University Hospitals Leuven, Belgium; Medizinische Hochschule Hannover, Germany; Centre Hospitalier Universitaire Limoges, France, and Hôpital Necker Paris, France). In all four clinical centers, protocol renal allograft biopsies were performed 3, 12 and sometimes 24 months after transplantation, in accordance with local practice, in addition to clinically indicated biopsies (biopsies at the time of graft dysfunction). In parallel, blood and urine samples were collected at the same times. All adult patients who had received a single renal allograft at these institutions and provided written informed consent for this study were eligible. This consent adheres to the Declaration of Istanbul. Ethics committee XI #13016, Paris, France for Necker Hospital gave ethical approval for this work. Ethics committee #S55598, Leuven, Belgium for Leuven Hospital gave ethical approval for this work. Ethics committee #6475, Hannover, Germany for Hannover Medical School gave ethical approval for this work. Ethics committee #DC-2010-1075, Limoges, France for Limoges Hospital gave ethical approval for this work. Recipients of combined transplantations were excluded. All transplantations were complement-dependent cytotoxicity cross-match negative. The study protocol was approved by institutional review boards and national regulatory agencies (where applicable) at each clinical center. The BIOMARGIN study was divided into three phases. Only data from the first exploratory phase are used in the present report. In this discovery phase, blood samples were used for an epigenome analysis (miRNA expression [E-MTAB-9595]^5^) and two transcriptome analyses (mRNA quantified by MicroArray [GSE129166]^1^, and bulk RNA sequencing [GSE175718]^3^). In parallel, biopsy samples were used for an epigenome analysis (miRNA expression [GSE179772]^4^) and a transcriptome analysis (mRNA quantified by MicroArray [GSE147089]^2^). With the exception of urinary mRNA, each dataset is publicly available, and details of RNA extractions and RNA expression analysis are extensively detailed in the corresponding reports.

### Urinary mRNA quantification

For mRNA quantified in urine, samples were centrifuged for 20 min at 2,000g at 4°C. The cell pellet was resuspended in 700µL of PBS and centrifuged for 5 min on a tabletop centrifuge at maximum speed. PBS was discarded and cells were resuspended in 500µL RLT buffer (Qiagen, Courtaboeuf, France) in a cryotube before being frozen at -80°C. MessengerRNA was extracted from the pellet using the RNeasy mini kit (Qiagen) and reverse transcribed into cDNA using TaqMan® reverse transcription reagents (Applied Biosystems). We used in-house designed oligonucleotide primers and fluorogenic probes to measure mRNA levels of ribosomal RNA 18S*, ACTA2*, *ENG, CD14, CD3E, CD46*, *CFB*, *CXCL13*, *CXCL9*, *CXCL10*, *IL2RA*, *CDH1, FN1*, *FOXP3*, *GZMB*, *GAPDH*, *HGF*, *PRF1, PSMB9*, *PSMB10*, *SLC12A1*, *TGFB1, TLR4* and *VIM* as detailed in Supplementary Table 1 or commercial assays (Thermofisher) for *CTSS* (Hs00175407_m1), *GNLY* (Hs00246266_m1), *FCGR3A* (Hs00275547_m1), *ISG20* (Hs00158122_m1), *KLRD1* (Hs00233844_m1), *MMP7* (Hs01042796_m1), *MMP9* (Hs00957562_m1), *NKG2D* (Hs00183683_m1), *NKG7* Hs01120688_g1), *RUNX3* (Hs00231709_m1), and *UPK1A* (Hs01086736_m1). PCR analysis was performed in two steps, a pre-amplification step as described previously^20^ (Veriti 96-Well Thermal Cycler, Applied Biosystems) followed by mRNA measurement with a Viia 7 Real-Time PCR system (Thermofisher). *GAPDH* was used as housekeeping gene^21^ for normalization using the delta Ct method. The normalized expression was log transformed before MOFA integration.

### Data processing and filtering

Both blood- and biopsy-derived miRNA expressions were determined as previously described^4,5^ using two small-nucleolar RNAs for normalization: RNU44 and RNU48. The normalized expressions were log transformed before MOFA integration. Blood- and biopsy-derived transcriptomes quantified by MicroArray were subjected to Robust Multichip Average (RMA) normalization as previously described^1,2^. These datasets were then curated and annotated using the Biological Interpretation Of Multiomics EXperiments (BIOMEX) workflow^22^ before MOFA integration. Raw counts corresponding to the blood-derived transcriptome quantified by bulk RNA sequencing were filtered to exclude weakly expressed genes (with <5 counts in 50% of the samples). After filtration, the data were subjected to normalization by variance stabilizing transformation (VST) using the R package DamiRseq (2.1.0)^23^ prior to MOFA integration.

### Multiple Omics Factor Analysis version 2

To integrate the six datasets, we used the R package MOFA2 (1.8.0). MOFA is an unsupervised machine learning method that identifies latent factors that capture biological sources of variability in multi-omics datasets. It should be noted that clinical covariates were not used to train the model. Data, model and learning options were left default. MOFA was run with 100 iterations in ‘slow’ convergence mode to ensure model convergence; the final model converged after 61 iterations. Interpretation of the factors is analogous to that of the principal components and the relationship between clinical covariates and MOFA factors was analyzed a posteriori.

### Clinicopathological diagnosis of acute rejection subtypes

Two kidney biopsy cores were obtained using a 14-gauge needle under sonographic guidance. One biopsy core was fixed in formalin and embedded in paraffin for standard histopathological assessment. Half of the second biopsy core was used for frozen sections and/or electron microscopy; the remaining half core was used for epigenome and transcriptome analyses. All biopsies were scored according to the internationally standardized Banff 2017 lesion scores^24^. The follow-up of anti-HLA antibodies and annotation of donor-specific antibodies (DSA) was systematically monitored in the histocompatibility laboratory referent of each inclusion center. A diagnostic label was awarded to each biopsy based on the presence and severity of these histological lesions and on the DSA status, in concordance with the Banff 2017 classification.

### Single-cell RNA-Sequencing (scRNA-Seq) validation

We reintegrated 46 publicly available scRNA-Seq datasets corresponding to kidney transplant biopsies or peripheral blood mononuclear cells (PBMCs) from patients presenting allograft rejection or not, whose raw data were downloaded from various repositories: E-MTAB-11450 ^25^, E-MTAB-12051 ^26^, GSE140989 ^27^, GSE145927 ^28^, GSE171374 ^29^, PRJNA974568 ^30^. Seurat R package (v5.0.1) was used to read, create and merge all raw counts matrices into a single object, which was subjected to the following QC parameters: number of features (genes) between 300 and 10,000 per cell and percent of total counts from mitochondrial genes, as defined by the prefix « MT-», below 10% for cells from PBMCs datasets, and below 25% for cells from kidney transplant biopsies datasets.

A total of 151,862 cells and 40,353 genes successfully passed QC. Data were log-normalized, scaled, the top 2000 variable features as well as the top 50 Principal Component (PC) dimensions were calculated prior to integration using Reciprocal Principal Component Analysis (RPCA) method, with GSM4339779 and pbmc5 as references and all other datasets as queries on a supercomputer (Mésocentre de calcul de Franche-Comté). Uniform Manifold Approximation and Projection (UMAP) dimensionality reduction was calculated from the top 50 PC dimensions, and a resolution of 0.47 was used for unsupervised clustering. scDblFinder R package (v1.16.0) was used for doublets discrimination (as defined by clusters composed of two or more different cells captured in a droplet and sequenced). Kidney-derived clusters annotation was made using already-described canonical markers^25–36^. PBMC-derived clusters automatic annotation was performed using SingleR (v2.4.1) and celldex (v1.12.0) R packages and MonacoImmuneData as reference dataset ^37^. Seurat, scCustomize ^38^ (v2.0.1) and RightSeuratTools ^39^ (v1.0.1) R packages were used for data visualization.

### Whole Transcriptomic external validations

Various external datasets were used to validate the top features explaining the factors determined by MOFA.

#### a) External validation of Factor 1 top-weighted genes

To validate the potential for discrimination between AMR and TCMR patients by the top-weighted Factor-1-related genes, an external blood-derived transcriptomic dataset, GSE120649^40^, was used. This dataset includes bulk RNAseq analysis of whole blood cells isolated from 6 patients with histologically verified AMR and 4 patients with histologically verified TCMR after kidney transplantation. In brief, the BIOMEX pipeline was used to determine differentially expressed genes between the two patient groups.

#### b) External validation of Factor 2 top-weighted genes

To validate the potential for discrimination between C4d negative biopsies and C4d postitive biopsies by the top-weighted Factor-2-related genes, an external kidney allograft-derived transcriptomic dataset, E-GEOD-38262^41^, was used. This dataset includes 92 microarray datasets corresponding to kidney transplant biopsies from patients divided into seven groups classified according to their histopathological scores, whose raw data (CEL files) were downloaded from ArrayExpress (https://www.ebi.ac.uk/biostudies/arrayexpress/). oligo (1.66.0), hugene10sttranscriptcluster.db (8.8.0), hugene10stprobeset.db (8.8.0) and pd.hugene.1.0.st.v1 (3.14.1) R packages were used to read raw data and normalize expression, as well as to convert genes names from Affymetrix HuGene 1.0 st Probe IDs into ENSEMBL IDs. The expression matrix was subsequently analyzed using the BIOMEX pipeline. Here we selected only samples of interest corresponding to biopsy transcriptomes showing minimal, diffuse or focal C4d deposition in the absence of glomerular disease (N=25) or absence of C4d and glomerular disease (N=17). After PCA analysis, 3 outlier samples were excluded (GSM937601, GSM937665, GSM937666) and genes differentially expressed between the 2 groups were calculated.

#### c) External validation of Factor 4 top-weighted genes

To validate the differential expression of top-weighted genes of Factor 4 according to ATG treatment, an external transcriptomic dataset GSE10040^42^, was used. This dataset includes a MicroArray analysis of PBMC isolated from healthy volunteers and in vitro treated with or without ATG (10μg/mL), whose raw data (CEL files) were downloaded from Gene expression Omnibus (https://www.ncbi.nlm.nih.gov/geo/query/acc.cgi?acc=GSE10040). affy (1.70.0), hgu133plus2.db (3.13.0), and limma (3.48.3) R packages were used to read raw data and normalize expression, as well as to annotate genes into ENSEMBL IDs. The expression matrix was subsequently analyzed using the BIOMEX pipeline. In brief, the BIOMEX pipeline was used to determine which genes were differentially expressed between the two experimental groups.

### Statistical analysis

We report descriptive statistics using mean and standard deviation (or median and interquartile range for skewed distributions) for continuous variables or numbers, and percentages for discrete variables, for the full cohort and for the rejection subgroups. We used R Studio (2023.06.1+524) and GraphPad Prism (v10; GraphPad Software, San Diego, CA, United States) for statistical analysis and data presentation. The volcano plots were constructed using the R packages ggplot2 (3.4.1) and ggrepel (0.9.3) and were used to annotate the genes of interest.

## Data availability

All data presented in this present study are derived from the BIOMARGN study and are already publicly available, with the exception of urinary gene expression, which can be shared upon reasonable request. Epigenome analyses were performed using the following datasets: blood-derived miRNA expression E-MTAB-9595 and biopsy-derived miRNA expression GSE179772. Blood-derived transcriptome analyses were performed using the following datasets: MicroArray-quantified mRNA GSE129166 and bulk RNA sequencing GSE175718. Biopsy-derived transcriptome analysis was performed using the following dataset: GSE147089.

## Code availability

Code for MOFA2 is available at https://biofam.github.io/MOFA2/.

## Acknowledgements

We thank the clinical centers of the BIOMARGIN consortium, the clinicians, surgeons, nursing staff and patients. Our special thanks go to Ricard Argelaguet, the first author of the original article describing MOFA, for his precious help with feature selection. We also thank Dr David Hildeman, Krishna Roskin and Babacar Ndao for their help with scRNA-Seq alignment.

## Author Contributions Statement

C.T., B.L. and M.N. conceived and designed the study. C.T. performed the MOFA analyses. A.V., C.T and B.L. generated, analysed and interpreted the single cell and bulk transcriptomic data. C.T, D.A., J.C., H.D.L., W.G., P.M., M.R., V.S., E.V.L. and M.N. included patients and collected and interpreted the clinical data. C.T., B.L. and M.N. wrote the paper with contribution from all co-authors.

