## Supplementary material for "Multiomic analyses uncover immunological signatures in kidney transplantation": Supplemetal Information

Fig.S1

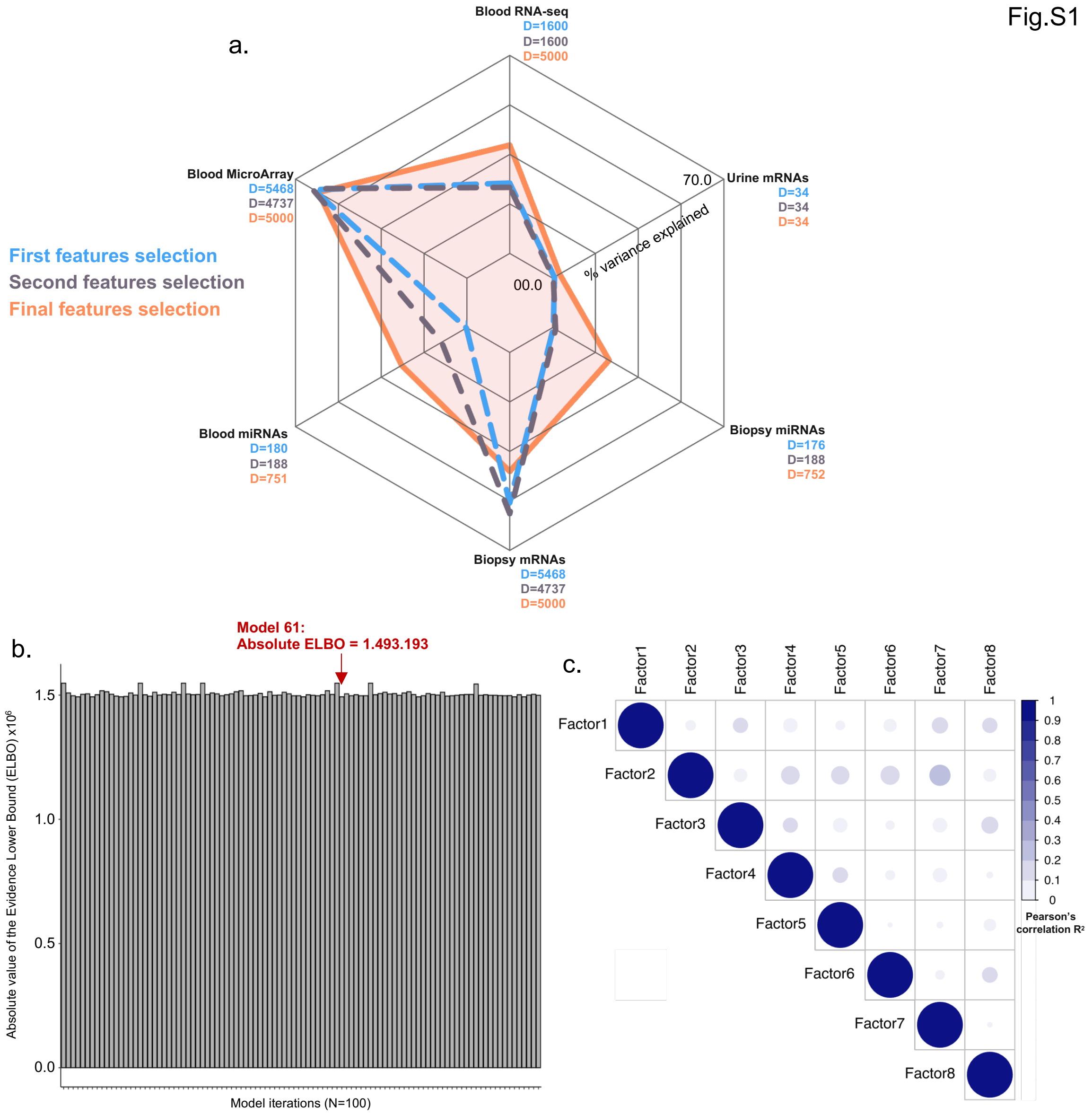


**Figure S1: MOFA model training and selection** (a) Percentages of variance explained in the 6 views according to the first 2 models tested and according to the final model selected. The number of features (D) per view and integrated in each model is indicated (b) Bargraph showing the absolute values of the Evidence Lower Bound (ELBO) of the 100 MOFA iterations. The Model 61 with the lowest ELBO value was selected (c) Pearson's correlation matrix analysis of the 8 latent factors discovered by the selected MOFA model.

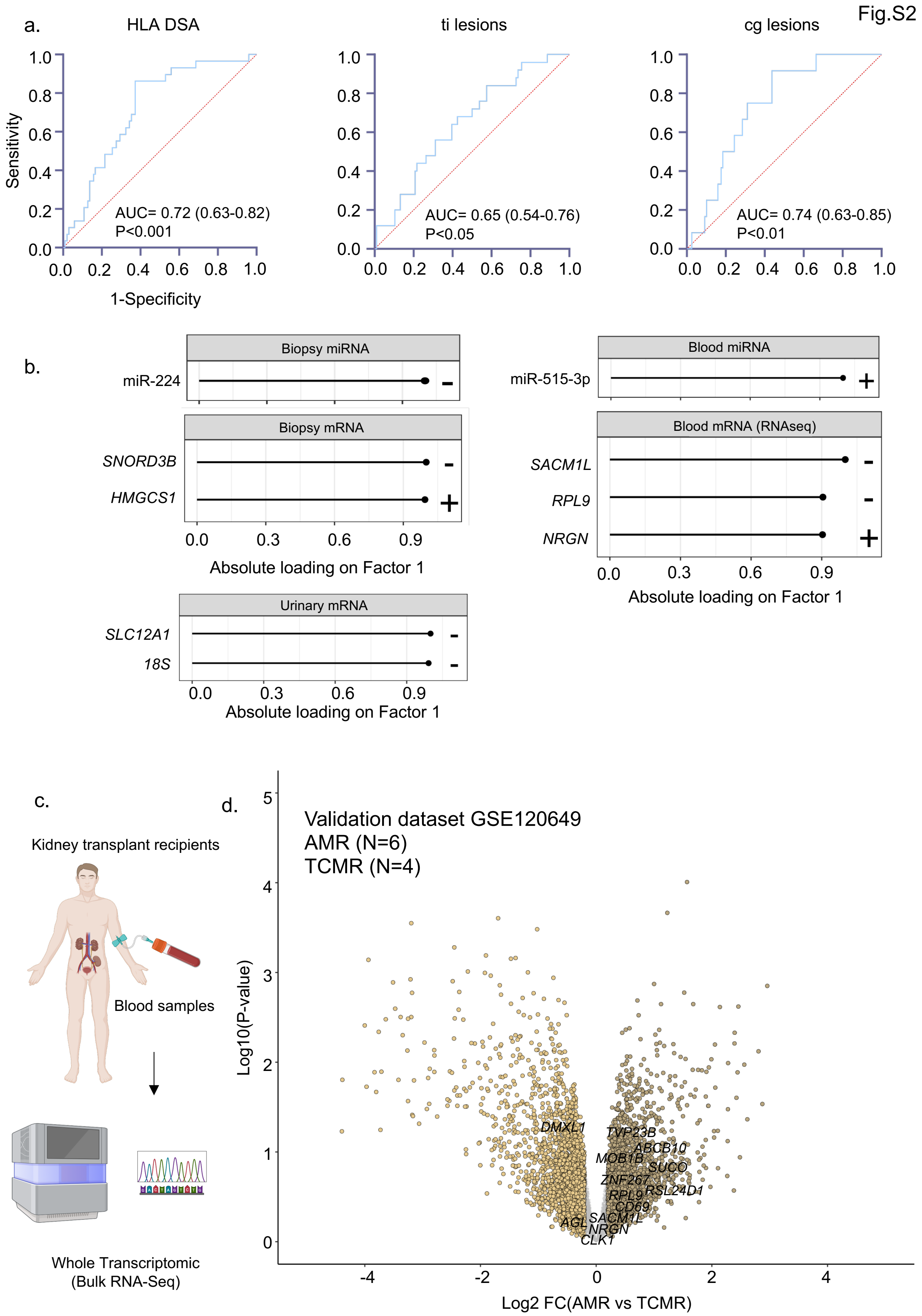


**Figure S2: Factor 1 interpretation (**a) ROC curves depicting the association between Factor 1 loading and clinical outcocmes (b) Lollipop graphs showing the top-weighted genes derived from the indicated datasets in latent Factor 1 (c-d) An external validation dataset GSE120649 was used to confirm the distribution of the top-weighted features explaining Factor 1 in blood samples from kidney transplanted patients showing AMR compared to TCMR. (c) Experimental scheme (d) Volcano plot depicting the differentially expressed genes in AMR compared TCMR patients' blood. The top-weighted genes in latent Factor 1 were annotated. Panel c was created using Biorender.com

Fig.S3

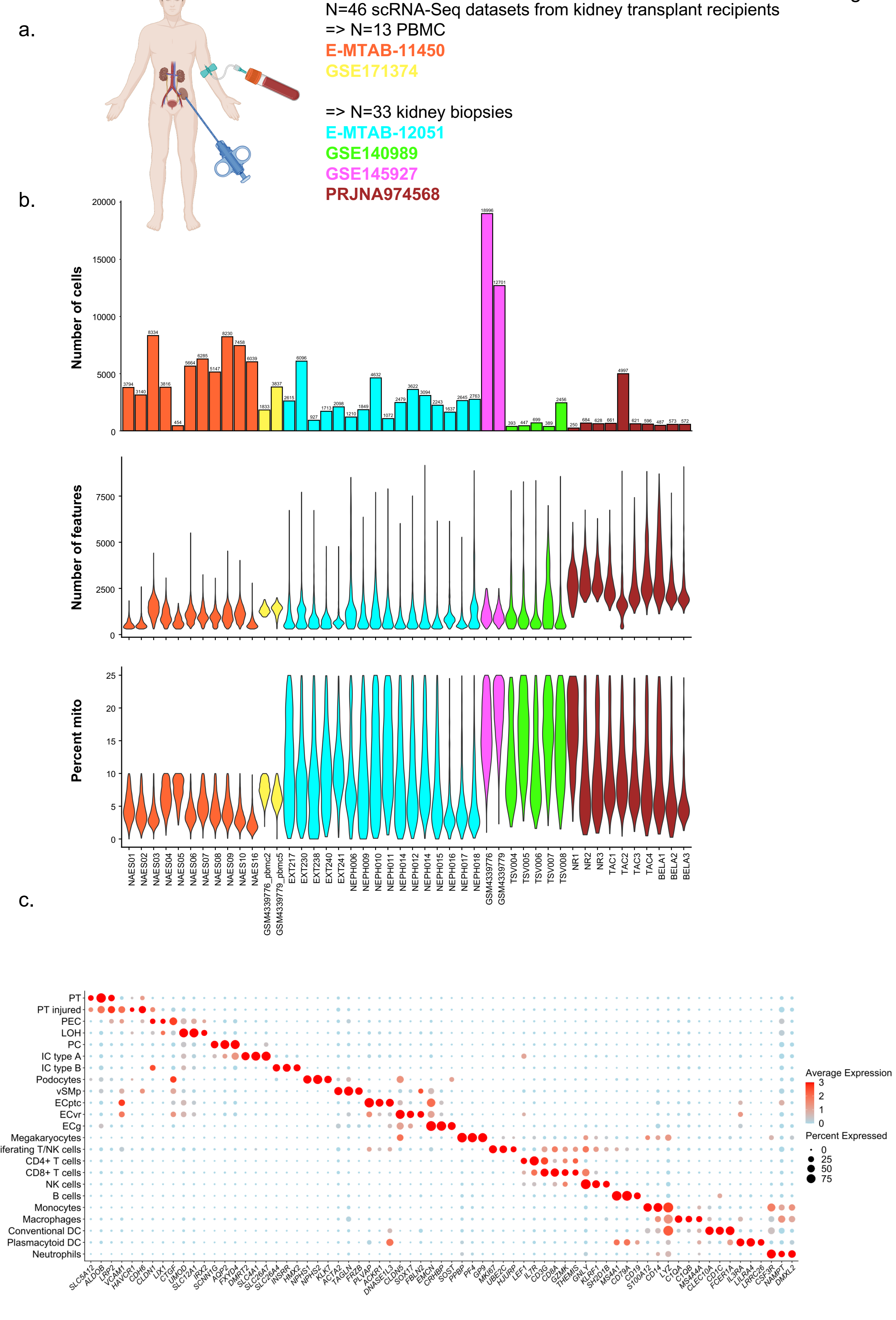


**Figure S3: Reintegration of scRNA-Seq datasets from kidney transplant recipients** (a) Indicated publicly available scRNA-Seq datasets were reintegrated. (b) Number of cells (top panel), of features (middle panel) and percentage of mitochondrial genes are indicated for each scRNA-Seq sample. (c) DotPlot showing the genes used for the clusters' annotation. Panel a was created using Biorender.com

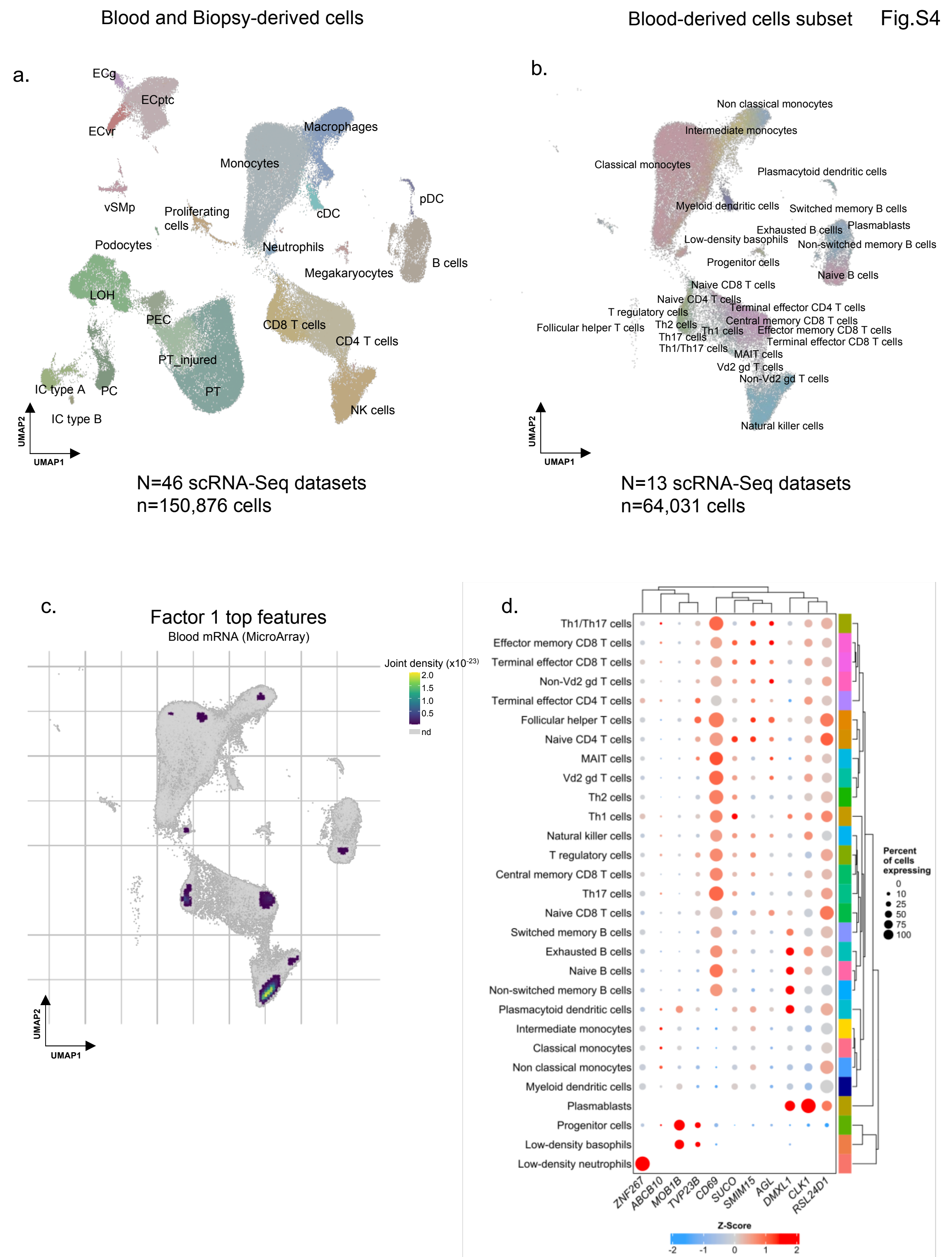


**Figure S4: Single-cell mapping of Factor 1 top features** (a) UMAP plot of the 150,876 integrated transcriptomes from kidney transplant recipients (b) UMAP plot showing the Single R automatic annotation of the blood-derived cells. (c) Joint density of the top features explaining Factor 1. (d) Dotplot showing the level of expression and percent of cells expressing the top features explaining Factor 1.

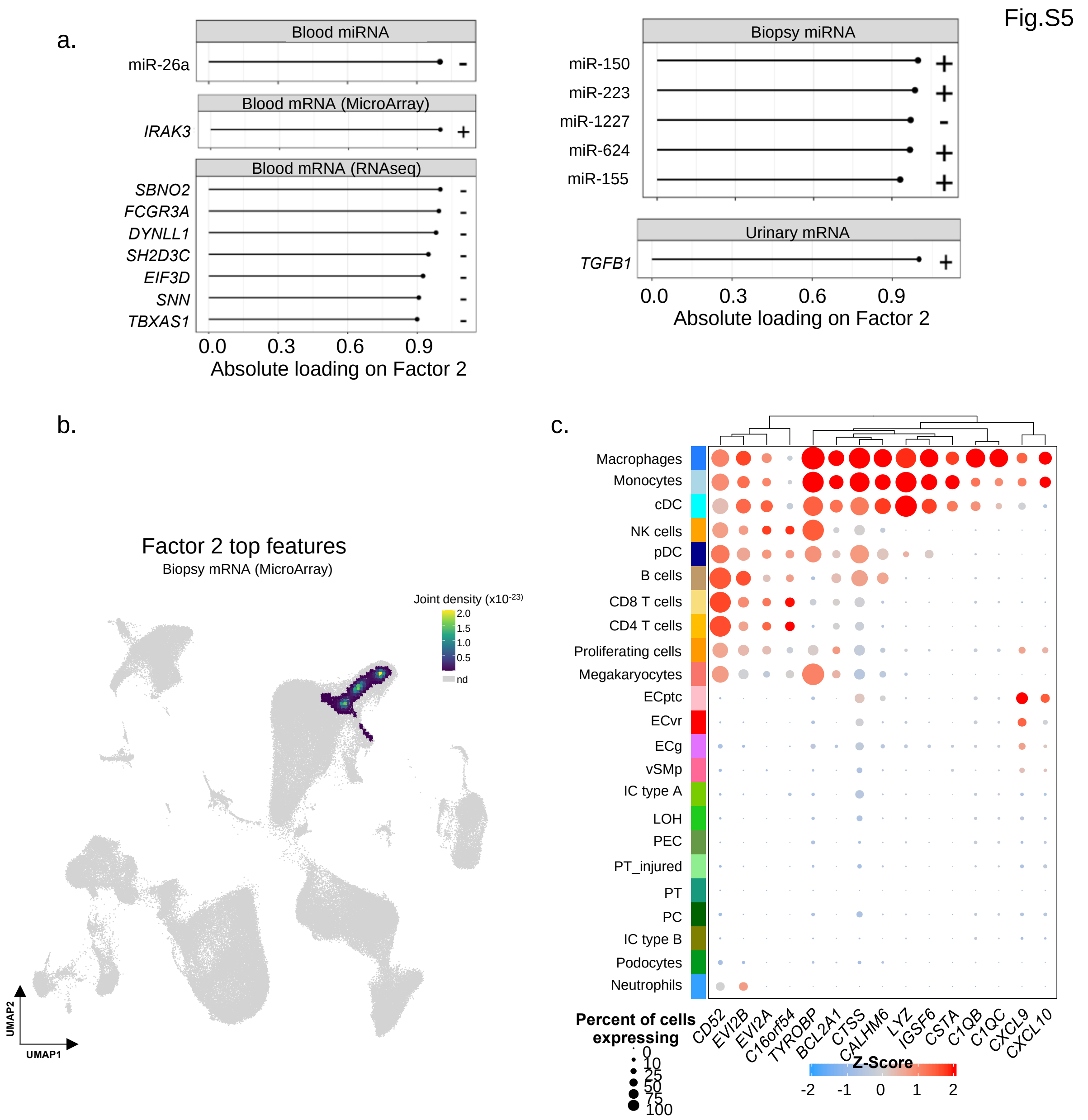


**Figure S5: Factor 2 characterization** (a) Lollipop graph showing the top features derived from the other views and explaining Factor 2. (b) Joint density of the top features explaining Factor 2. (d) Dotplot showing the level of expression and percent of cells expressing the top features explaining Factor 2.

Fig.S6

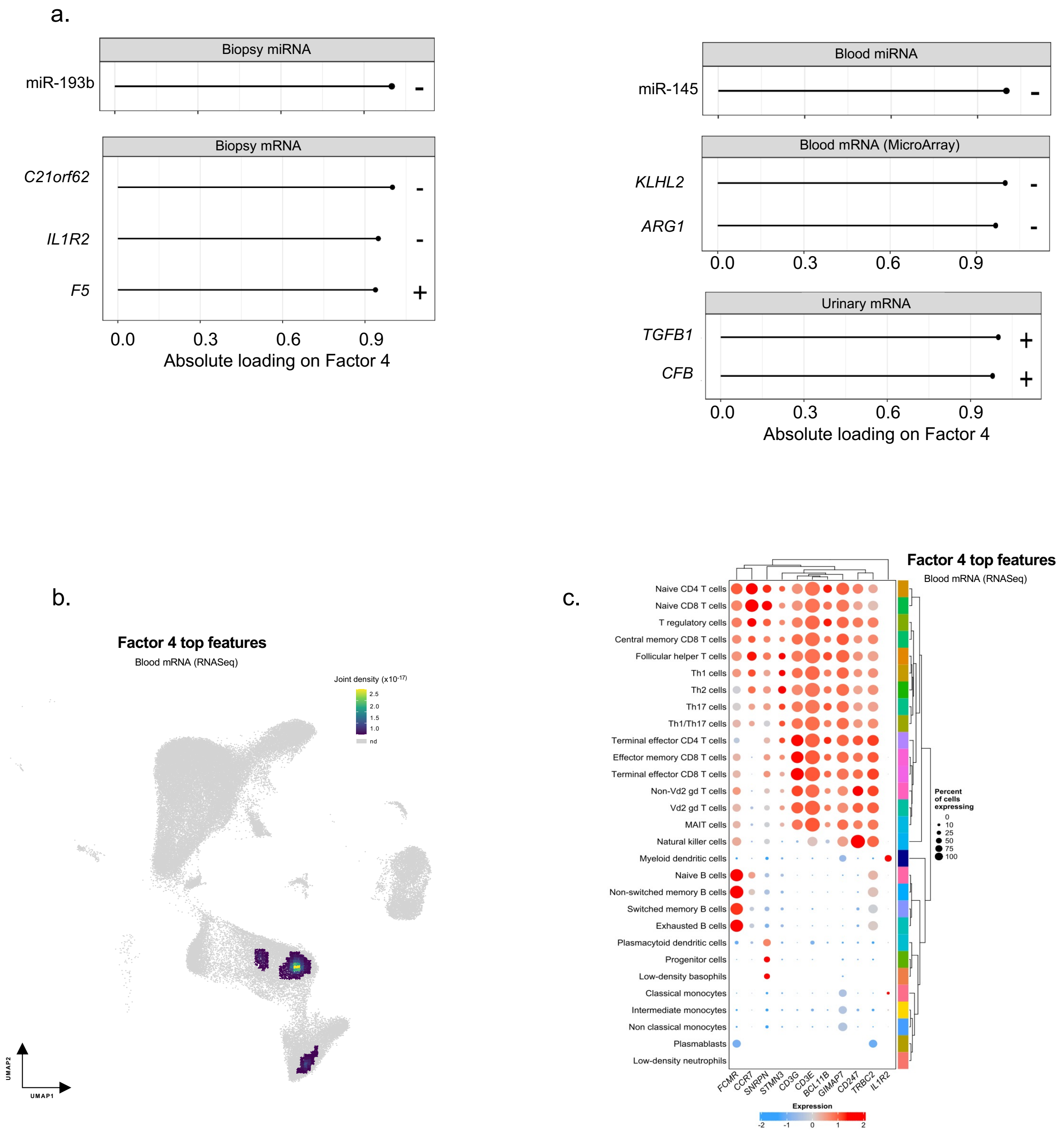


**Figure S6: Factor 4 characterization** (a) Lollipop graph showing the top features derived from the other views and explaining Factor 4. (b) Joint density of the top features explaining Factor 4. (d) Dotplot showing the level of expression and percent of cells expressing the top features explaining Factor 4.

| Gene | Ensembl transcript annotation | Sequence | Probes |
|---|---|---|---|
| *HGF* | ENST00000222390.11 | Sense: 5' CAAATGTCAGCCCTGGAGTTC 3'<br>Antisense: 5' CTGTAGGTCTTTACCCCGATAGCT 3' | 5’ FAM ATGATACCACACGAACACAGCTTTTTGCC TAMARA 3’ |
| *ACTA2* | ENST00000224784.10 | Sense: 5’ TGGGACGACATGGAAAAGATC 3’<br>Antisense: 5’ CAGGGTGGGATGCTCTTCAG 3’ | 5’ FAM CCACTCTTTCTACAATGAGCTTCGTGTTGCC TAMRA 3’ |
| *FN1* | ENST00000354785.11 | Sense: 5' GAAAGTACACCTGTTGTCATTCAACA 3'<br>Antisense: 5' ACCTTCACGTCTGTCACTTCCA 3' | 5' FAM CCACTGGCACCCCACGCTCA TAMRA 3' |
| *ENG* | ENST00000373203.9 | Sense: 5’ CAGCCTCAGCCCCACAAGT 3’<br>Antisense: 5’ GGCCCACAGGCTGAAGGT 3’ | 5’ FAM TTGCAGAAACAGTCCATT MGB 3’ |
| *TGFB1* | ENST00000221930.6 | Sense: 5' GCGTGCTAATGGTGGAAACC 3'<br>Antisense: 5' CGGAGCTCTGATGTGTTGAAGA 3' | 5’ FAM ACAACGAAATCTATGACAAGTTCAAGCAGAGTACACA TAMRA 3’ |
| *GZMB* | ENST00000216341.1 | Sense: 5' GCGAATCTGACTTACGCCATTATT 3'<br>Antisense: 5' CAAGAGGGCCTCCAGAGTCC 3' | 5’ FAM CCCACGCACAACTCAATGGTACTGTCG TAMRA 3’ |
| *VIM* | ENST00000224237.9 | Sense: 5’TCAGAGAGAGGAAGCCGAAAAC 3’<br>Antisense: 5’ CCAGAGACGCATTGTCAACATC 3’ | 5’ FAM CCCTGCAATCTTTCAGAC MGB 3’ |
| *CD3E* | ENST00000361763.9 | Sense: 5' AAGAAATGGGTGGTATTACACAGACA 3'<br>Antisense: 5' TGCCATAGTATTTCAGATCCAGGAT 3' | 5' FAM CCATCTCTGGAACCACAGTAATATTGACATGCC TAMRA 3' |
| *CD46* | ENST00000367042.6 | Sense: 5' GATCGGAATCATACATGGCTACCT 3'<br>Antisense: 5' GGCCATTTAAAGGATCCCGTATA 3' | 5’ FAM CTCAGATGACGCCTGTTATAGAGAAACATGTCCA TAMRA 3’ |
| *CXCL10* | ENST00000306602.3 | Sense: 5' TGTCCACGTGTTGAGATCATTG 3'<br>Antisense: 5' GGCCTTCGATTCTGGATTCA 3' | 5’ FAM TACAATGAAAAAGAAGGGTGAGAA MGB 3’ |
| *SLC12A1* | ENST00000380993.8 | Sense: 5’ TCACGAGCAACTCGCAAAGA 3’<br>Antisense: 5’ TCCCATCACCGTTAGCAACTC 3’ | 5’ FAM TGTGGCAGTCACCCCAAGTTCAGC TAMRA 3’ |
| *IL2RA* | ENST00000379959.8 | Sense: 5’ GACTGCTCACGTTCATCATGGT 3’<br>Antisense: 5’ AATGTGGCGTGTGGGATCTC 3’ | 5' FAM AGAGCTCTGTGACGATGACCCGCC TAMRA 3' |
| *FOXP3* | ENST00000684155.1 | Sense: 5’ GAGAAGCTGAGTGCCATGCA 3’<br>Antisense: 5’ GGAGCCCTTGTCGGATGAT 3’ | 5’ FAM TGCCATTTTCCCAGCCAGGTGG TAMRA3’ |
| *CDH1* | | Sense: 5' TGAGTGTCCCCCGGTATCTTC 3'<br>Antisense: 5' CAGCCGCTTTCAGATTTTCAT 3' | 5' FAM CCTGCCAATCCCGATGAAATTGGAAAT TAMRA 3' |
| *PRF1* | ENST00000373209 | Sense: 5' GGACCAGTACAGCTTCAGCACTG 3'<br>Antisense: 5' GCCCTCTTGAAGTCAGGGTG 3' | 5' FAM TGCCGCTTCTACAGTTTCCATGTGGTACAC TAMRA 3' |
| *CXCL9* | ENST00000264888.6 | Sense: 5' CTTTTCCTCTTGGGCATCATCT 3'<br>Antisense: 5' AGGAACAGCGACCCTTTCTCA 3' | 5’ FAM TACTGGGGTTCCTTGCACTCCAATCAGA TAMRA 3’ |
| *TLR4* | ENST00000355622 | Sense: 5’ CATGGCCTTCCTCTCCTGC 3’<br>Antisense: 5’ GAAATTCAGCTCCATGCATTGA 3’ | 5’ FAM AGGAACCACCTCCACGCAGGGCTTAMRA 3’ |
| *CD14* | ENST00000303014 | Sense: 5’ GCTGTGTAGAAAGAAGCTAAAGCACTT 3’<br>Antisense: 5’ TGGCGTGGTCGCAGAGA 3’ | 5’ FAM CTTATCGACCATGGAGCGCGCGT TAMRA 3’ |
| *CFB* | ENST00000375455 | Sense: 5' TGGCGGCCCCTTGATAGT 3'<br>Antisense: 5' CCCAGCTGATTACACCAACTTG 3' | 5’ FAM CACAAGAGAAGTCGTTTCA MGB 3’ |

| | | | |
|---|---|---|---|
| *CXCL13* | ENST00000286758 | Sense: 5’ CCCGTGGGAATGGTTGTC 3’<br>Antisense: 5’ GGGTCCACACACACAATTGACT 3’ | 5’ FAM ATCATAGTCTGGAAGAAGAA MGB 3’ |
| *PSMB10* | ENST00000682537.1 | Sense: 5' AGAGCTGCGAGAAGATCCACTT 3’<br>Antisense: 5’ CTCCAGCCCCACAGCAGTA 3’ | 5’ FAM ATCGCCCCCAAAAT MGB 3’ |
| *COL1A1* | ENST00000507689 | Sense: 5' CCAGAAGAACTGGTACATCAGCAA3'<br>Antisense: 5' CGCCATACTCGAACTGGAATC3' | 5’ FAM ACAAGAGGCATGTCTGG MGB 3’ |
| *GAPDH* | ENST00000229239 | Sense: 5’ CCACATCGCTCAGACACCAT 3’<br>Antisense: 5’ TGACCAGGCGCCCAATA 3’ | 5’ FAM AGTCAACGGATTTGGTC MGB 3’ |
| 18S rRNA | NA | Sense: 5' GCCCGAAGCGTTTACTTTGA 3'<br>Antisense: 5' TCCATTATTCCTAGCTGCGGTATC 3' | 5’ FAM AAAGCAGGCCCGAGCCGCC TAMRA 3’ |

### Supplemental Table 1 Primers for urinary RT-qPCR

| Variables | All samples *N*=131 | AMR *N*=31 | IFTA *N*=38 | Normal *N*=45 | TCMR *N*=17 | *P*-value |
|---|---|---|---|---|---|---|
| Sex (men), *N (%)* | 79 (60.3) | 16 (51.6) | 22 (57.9) | 31 (68.9) | 10 (58.8) | 0.4805 |
| Age at transplantation (yr), *mean±SD* | 50±14 | 46±14 | 50±14 | 50±14 | 57±14 | 0.0748 |
| Donor age (yr), *mean±SD* | 51±16 | 47±15 | 54±15 | 47±16 | 58±16 | **0.0484** |
| Living donor, *N (%)* | 25 (19.1) | 5 (16.1) | 3 (7.9) | 15 (33.3) | 2 (11.8) | **0.0248** |
| Retransplantation, *N (%)* | 20 (15.3) | 11 (35.5) | 3 (7.9) | 4 (8.9) | 2 (11.8) | **0.0336** |
| CIT (h), *mean±SD ** | 13±7 | 16±8 | 14±7 | 10±8 | 14±5 | **0.0136** |
| Delayed graft function, *N (%)* | 21 (16.0) | 8 (25.8) | 6 (15.8) | 5 (11.1) | 2 (11.8) | 0.3305 |
| Post-transplant time (months), *median (IQR)* | 12.0 (34.2) | 11.8 (42.8) | 24.7 (58.1) | 11.6 (10.0) | 3.5 (10.7) | **<0.0001** |
| Preformed HLA-DSA, *N (%)* | 29 (23.2) | 18 (58.1) | 2 (5.3) | 5 (11.1) | 4 (23.5) | **<0.0001** |
| **Cause of ESRD** | | | | | | |
| Glomerulonephritis, *N (%)* | 34 (26.0) | 8 (25.8) | 8 (21.1) | 12 (26.7) | 6 (35.3) | 0.7389 |
| Diabetes, *N (%)* | 10 (7.6) | 3 (9.7) | 5 (13.2) | 2 (4.4) | 0 (0.0) | 0.3189 |
| Cystic/hereditary/congenital, *N (%)* | 36 (27.5) | 11 (35.5) | 9 (23.7) | 13 (28.9) | 3 (17.6) | 0.5583 |
| Secondary glomerulonephritis, *N (%)* | 6 (4.6) | 3 (2.3) | 0 (0.0) | 2 (4.4) | 1 (5.9) | 0.2390 |
| Hypertension, *N (%)* | 4 (3.0) | 0 (0.0) | 2 (5.3) | 1 (2.2) | 1 (5.9) | 0.5120 |
| Interstitial nephritis, *N (%)* | 9 (6.9) | 1 (3.2) | 4 (10.5) | 4 (8.9) | 0 (0.0) | 0.4941 |
| Miscellaneous conditions, *N (%)* | 9 (6.9) | 1 (3.2) | 1 (2.6) | 3 (6.7) | 4 (23.5) | 0.0575 |
| Uncertain etiology, *N (%)* | 23 (17.6) | 4 (12.9) | 9 (23.7) | 8 (17.8) | 2 (11.8) | 0.6654 |
| **Immunosuppression protocol** | | | | | | |
| Induction therapy, *N (%)* | 100 (77.0) | 30 (96.8) | 25 (65.8) | 34 (75.6) | 11 (64.7) | **0.0049** |
| Basiliximab, *N (%)* | 75 (57.7)) | 18 (58.1) | 19 (50.0) | 27 (60.0) | 11 (64.7) | 0.7290 |
| ATG, *N (%)* | 22 (16.9) | 11 (35.5) | 4 (10.5) | 7 (15.6) | 0 (0.0) | **0.0078** |
| CNI: Cyclosporin, *N (%)* | 23 (17.6) | 5 (16.1) | 11 (28.9) | 4 (8.9) | 3 (17.6) | 0.1252 |
| Tacrolimus, *N (%)* | 108 (82.4) | 26 (83.9) | 27 (71.1) | 41 (91.1) | 14 (82.4) | 0.1252 |
| Mycophenolic acid, *N (%)* | 126 (96.2) | 29 (93.5) | 35 (92.1) | 45 (100.0) | 17 (100.0) | 0.1493 |
| mTOR inhibitors, *N (%)* | 1 (0.8) | 0 (0.0) | 1 (2.6) | 0 (0.0) | 0 (0.0) | 0.6565 |
| Steroids, *N (%)* | 131 (100.0) | 31 (100) | 38 (100) | 45 (100.0) | 17 (100.0) | >0.9999 |

**Supplemental Table 2  Patients characteristic**

| Variables | Factor1>median *N*=65 | Factor1≤median *N*=66 | *P*-value |
|---|---|---|---|
| Sex (men), *N (%)* | 44 (67.7) | 35 (53.0) | 0.0864 |
| Age at transplantation (yr), *mean±SD* | 52±*14* | 48±*14* | 0.0902 |
| Donor age (yr), *mean±SD* | 51±*14* | 50±*17* | 0.8659 |
| Living donor, *N (%)* | 11 (16.9) | 14 (21.2) | 0.5322 |
| Retransplantation, *N (%)* | 8 (12.3) | 12 (18.2) | 0.3500 |
| CIT (h), *mean±SD ** | 13.2±7.3 | 12.8±7.6 | 0.9753 |
| Delayed graft function, *N (%)* | 12 (18.4) | 9 (13.6) | 0.4517 |
| Post-transplant time (months), *median (IQR)* | 12 (32) | 12 (41) | 0.6207 |
| Preformed HLA-DSA, *N (%)* | 4 (6.1) | 25 (37.9) | **<0.0001** |
| **Histological Banff lesions** | | | |
| g | 8 (12.3) | 22 (33.3) | **0.0042** |
| ptc | 11 (16.9) | 24 (36.4) | **0.0119** |
| cg | 1 (1.5) | 13 (19.7) | **0.0010** |
| i | 18 (28.6) | 10 (15.1) | 0.0646 |
| t | 18 (30.0) | 13 (19.7) | 0.1799 |
| ti | 31 (47.7) | 22 (33.3) | 0.0941 |
| v | 2 (3.3) | 3 (4.7) | >0.9999 |
| ct | 46 (71.9) | 38 (57.6) | 0.0883 |
| ci | 38 (59.4) | 34 (51.5) | 0.3674 |
| ah | 35 (54.7) | 30 (45.4) | 0.2925 |
| cv | 35 (56.4) | 30 (46.1) | 0.2822 |
| **Phenotype** | | | |
| AMR | 9 (13.9) | 22 (33.3) | **0.0087** |
| IFTA | 20 (15.3) | 18 (13.7) | 0.6593 |
| Normal | 24 (36.9) | 21 (31.8) | 0.5384 |
| TCMR | 12 (9.2) | 5 (3.8) | 0.0736 |

**Supplemental Table 3: Patients characteristic according to Factor 1 median**

| Variables | Factor2>median *N*=65 | Factor2≤median *N*=66 | *P*-value |
|---|---|---|---|
| Sex (men), *N (%)* | 35 (53.8) | 44 (66.7) | 0.1337 |
| Age at transplantation (yr), *mean±SD* | 49±15 | 51±13 | 0.548 |
| Donor age (yr), *mean±SD* | 51±17 | 50±14 | 0.5255 |
| Living donor, *N (%)* | 10 (15.4) | 15 (22.7) | 0.2849 |
| Retransplantation, *N (%)* | 13 (20.0) | 7 (10.6) | 0.1350 |
| CIT (h), *mean±SD ** | 13.7±7.2 | 12.4±7.6 | 0.3178 |
| Delayed graft function, *N (%)* | 11 (16.9) | 10 (15.1) | 0.7823 |
| Post-transplant time (months), *median (IQR)* | 12 (35) | 12 (32) | 0.3645 |
| Preformed HLA-DSA, *N (%)* | 15 (23.1) | 14 (21.2) | 0.8357 |
| C4d positivity | 11 (19.3) | 6 (9.7) | 0.1341 |
| **Phenotype** | | | |
| AMR | 22 (40.0) | 9 (13.6) | **0.0009** |
| IFTA | 20 (30.7) | 18 (22.3) | 0.6593 |
| Normal | 12 (21.8) | 33 (50.0) | **0.0014** |
| TCMR | 11 (16.9) | 6 (10.7) | 0.4335 |

**Supplemental Table 4 Patients characteristic according to Factor 2 median**

| Variables | Factor4>median *N*=65 | Factor4≤median *N*=66 | *P*-value |
|---|---|---|---|
| Sex (men), *N (%)* | 37 (56.9) | 42 (63.6) | 0.4323 |
| Age at transplantation (yr), *mean±SD* | 47±15 | 53±13 | 0.0613 |
| Donor age (yr), *mean±SD* | 48±17 | 53±14 | **0.0429** |
| Living donor, *N (%)* | 7 (10.7) | 18 (27.3) | **0.0162** |
| Retransplantation, *N (%)* | 14 (21.5) | 6 (9.1) | **0.0476** |
| CIT (h), *mean±SD ** | 13.6±6.1 | 12.4±8.5 | 0.3429 |
| Delayed graft function, *N (%)* | 7 (10.7) | 14 (21.2) | 0.1033 |
| Post-transplant time (months), *median (IQR)* | 14 (58) | 12 (34) | 0.0683 |
| Preformed HLA-DSA, *N (%)* | 11 (16.9) | 18 (27.3) | 0.1537 |
| **Phenotype** | | | |
| AMR | 13 (20.0) | 18 (27.3) | 0.3275 |
| IFTA | 20 (30.8) | 18 (27.3) | 0.6593 |
| Normal | 22 (33.8) | 23 (34.8) | 0.9039 |
| TCMR | 10 (7.6) | 7 (5.3) | 0.4475 |
| **Cause of ESRD** | | | |
| Glomerulonephritis, *N (%)* | 21 (32.3) | 13 (19.7) | 0.0997 |
| Diabetes, *N (%)* | 4 (6.1) | 6 (9.1) | 0.7441 |
| Cystic/hereditary/congenital, *N (%)* | 18 (27.7) | 18 (27.3) | 0.9571 |
| Secondary glomerulonephritis, *N (%)* | 3 (4.6) | 3 (4.5) | >0.9999 |
| Hypertension, *N (%)* | 0 (0.0) | 4 (6.1) | 0.1193 |
| Interstitial nephritis, *N (%)* | 5 (7.7) | 4 (6.1) | 0.7441 |
| Miscellaneous conditions, *N (%)* | 4 (6.1) | 5 (7.6) | >0.9999 |
| Uncertain etiology, *N (%)* | 10 (15.4) | 13 (19.7) | 0.5166 |
| **Immunosuppression protocol** | | | |
| Induction therapy, *N (%)* | 45 (69.2) | 55 (83.3) | 0.0576 |
| Basiliximab, *N (%)* | 39 (60.0) | 36 (54.5) | 0.5281 |
| ATG, *N (%)* | 4 (6.1) | 18 (27.3) | **0.0012** |
| CNI: Cyclosporin, *N (%)* | 8 (12.3) | 15 (22.7) | 0.1171 |
| Tacrolimus, *N (%)* | 57 (87.7) | 51 (77.3) | 0.1171 |
| Mycophenolic acid, *N (%)* | 61 (93.8) | 65 (98.5) | 0.2079 |
| mTOR inhibitors, *N (%)* | 0 (0.0) | 1 (0.8) | >0.9999 |
| Steroids, *N (%)* | 65 (49.6) | 66 (50.38) | >0.9999 |

**Supplemental Table 5 Patients characteristic according to Factor 4 median**
